# Assessing Supervised Natural Language Processing (NLP) Classification of Violent Death Narratives: Development and Assessment of a Compact Large Language Model (LLM) Approach

**DOI:** 10.1101/2025.01.16.25320680

**Authors:** Susan T. Parker

**Affiliations:** Research Assistant Professor Northwestern University Feinberg School of Medicine

**Keywords:** natural language processing, violence, computer simulation, homicide

## Abstract

**Objective:** The recent availability of law enforcement and coroner/medical examiner reports for nearly every violent death in the US expands the potential for natural language processing (NLP) research into violence. The objective of this work is to assess applications of supervised NLP to unstructured narrative data in the National Violent Death Reporting System (NVDRS).

**Materials and Methods:** This analysis applied distilBERT, a compact LLM, to unstructured narrative data to simulate the impacts of pre-processing, volume and composition of training data on model performance, evaluated by F1-scores, precision, recall and the false negative rate. Model performance was evaluated for bias by race, ethnicity, and sex by comparing F1-scores across subgroups.

**Results:** A minimum training set of 1,500 cases was necessary to achieve an F1-score of 0.6 and a false negative rate of .01-.05 with a compact LLM. Replacement of domain-specific jargon improved model performance while oversampling positive class cases to address class imbalance did not substantially improve F1 scores. Between racial and ethnic groups, F1-score disparities ranged from 0.2 to 0.25, and between male and female victims differences ranged from 0.12 to 0.2.

**Discussion:** Findings demonstrate that compact LLMs with sufficient training data can be applied to supervised NLP tasks to events with class imbalance in NVDRS unstructured police and coroner/medical examiner reports.

**Conclusion:** Simulations of supervised text classification across the model-fitting process of pre-processing and training a compact LLM informed NLP applications to unstructured death narrative data.

## INTRODUCTION

Violent injuries are among the leading causes of death in the United States for individuals under the age of 44, and are leading causes for young people aged 10-34.^1^ The most comprehensive and detailed source of data on violent deaths in the United States is the National Violent Death Reporting System (NVDRS), aggregating information from death certificates, coroner/medical examiner reports, and law enforcement reports to characterize violent deaths.^2^ Researchers have used structured data from NVDRS extensively to characterize the epidemiology of violent deaths including homicides,^3–6^ suicides,^7–10^ and those that result from legal intervention (police shootings).^11^

While NVDRS has been widely used for its structured data, which captures information such as victim characteristics, weapons, circumstances, and suspect information,^12^ far less attention has been given to the vast amounts of unstructured text data embedded within the narrative reports. Narratives provide rich details about the incident not necessarily captured in structured variables, such as nuanced descriptions of precipitating events and other contextual factors that are difficult to quantify.

Despite the rich information these narratives provide, use of NVDRS narratives in research has been limited. Studies using narratives have, with few exceptions,^13,14^ mostly relied on labor-intensive manual review and qualitative coding methods to analyze narrative content.^15–19^ Machine learning techniques designed to analyze unstructured text, known as natural language processing (NLP), have the potential to enable researchers to use NVDRS narrative data more efficiently, and thus to take on research questions that might otherwise be resource intensive. Applications of NLP to a related text narrative type, clinical notes from medical providers, have identified patient self-harm^20–25^ and violence-related^26–29^ outcomes. Developing applications of NLP to NVDRS is particularly important because the volume of NVDRS data will substantially increase over time. NVDRS has gathered data on over 500,000 deaths since 2003 and will grow by approximately 100,000 records annually moving forward as additional states and counties participate.

Although large language models (LLMs) have generally performed better than other NLP approaches to narrative data in medical informatics domains, few applications of LLMs to NVDRS exist.^13,14^ In part, researchers and practitioners may face particular challenges applying LLMs to NVDRS. One important challenge is that many outcomes of interest are likely to be infrequent or rare events that can present classification challenges due to sparse information about the outcome.^30–34^ Further, NVDRS narratives are composed of police and coroner reports which contain domain-specific language, or jargon, such as use of International Classification of Disease (ICD) codes.^35–40^ NVDRS data restrictions on sensitive data do not permit narratives to be stored in the cloud thus limiting access to computing resources that are often used to train or fine-tune LLMs. Fourth, researchers documented racial disparities in narratives alongside gendered text differences in NVDRS.^41–44^ Narratives involving victims from marginalized populations tend to be significantly shorter in length and are more likely to be missing altogether. These differences in data quality may result in models that generate predictions with similar patterns of subgroup bias.

To address these challenges, this paper conducts simulations of supervised text classification that span the machine learning pipeline, from data preprocessing and model training to the evaluation of predictions for potential racial or gender bias. Text classification outcomes with class imbalance were selected, as this setting is likely of most use to NVDRS applications, and models were fit using a compact LLM to reflect settings where computing resources are limited. By conducting simulations, this analysis aims to inform future applications of supervised classification using LLMs to NVDRS by establishing concrete benchmarks for understanding training data quantity, pre-processing needs, and to what extent NLP results in predictions reflecting existing racial and/or gender bias in narratives.

## METHODS

### Data

This analysis used violent death records from NVDRS data from 2015-2020. The National Violent Death Reporting System gathers information about violent deaths including homicides, suicides, and deaths caused by law enforcement. NVDRS combines data from death certificates, coroner/medical reports and law enforcement reports, providing context about violent deaths including information about mental health conditions, toxicology results, and other circumstances in addition to detail about victim characteristics. Trained abstractors code information about violent deaths into the over 600 variables that comprise the NVDRS surveillance system.^12^

To obtain labeled outcomes for use as target outcomes in simulations, this analysis constructed measures from existing coded NVDRS variables that abstractors label. Because a substantial proportion of coded NVDRS fields group together case outcomes that are negative with those that are not known, this analysis instead relied on multinomial fields or combined separate NVDRS coded variables to obtain target outcomes for simulations. For instance, for case outcomes such a mental health crisis or drug involvement, outcomes are coded as “Yes” or as “No, Not Available, Unknown,” which would not constitute a labeled outcome.

These constructed outcomes include four binary outcomes likely to be recorded accurately when known. The first outcome is whether or not a homicide is a legal intervention homicide, meaning the shooter was a law enforcement officer. Literature suggests that these homicides are well-recorded in NVDRS and less subject to noisy labeling or measurement error.^11^ The second outcome is whether or not a homicide is classified as a driveby shooting. The third outcome is whether a homicide occurred at home or not, and the fourth outcome is whether or not additional victims were non fatally shot in the course of a homicide event. we constrain the sample to where the weapon type is listed as firearm and the abstractor manner of death is a homicide. Taken together, these outcomes represent a range of language complexity and frequency less subject to label noise by constructing outcomes.

### Statistical Analysis

This analysis compared model performance across four configurations of training data and text composition using a compact large language model (LLM). The configurations examined included pre-processing of text data as well as the amount and composition of the training data. Specifically, the analysis first varied the amount of training data that the model was fitted on to inform how much randomly sampled training data must be annotated to train a LLM to predict NVDRS outcomes. Second, because positive class cases were often infrequent, the analysis simulated oversampling of positive class cases in training data. Specifically, oversampling included a larger proportion of additional positive class cases, holding the negative class cases constant, to inform what composition of training data was most effective to include as training data.

This analysis additionally simulated different preprocessing techniques for unstructured text data. NVDRS text may be domain-specific as it comprises police and coroner reports which use both jargon and abbreviation. To simulate the impacts of clarifying common abbreviations, this analysis replaced NVDRS abbreviations with unabbreviated text. For example, often when NVDRS abstractors referred to victims and suspects in the report narratives, the abbreviations “v” for victim and “s” for suspect appeared rather than the full word. Abbreviations referring to victims, suspects, police, and gunshot wounds were replaced (see Appendix Table 1).

**Table 1:** Sample Descriptive Statistics, Characteristics by Outcome.

|  | Drive-by Shooting |  |  | Legal Intervention |  |  | Number Nonfatally Shot |  |  | Victim Injured at Home |  |  |
| --- | --- | --- | --- | --- | --- | --- | --- | --- | --- | --- | --- | --- |
| Target outcome | Total | Neg | Pos | Total | Neg | Pos | Total | Neg | Pos | Total | Neg | Pos |
| **Count N, (% of category)** | 71,708<br>(100) | 65133<br>(91.7) | 6575<br>(9.2) | 76,197<br>(100) | 71,708<br>(94.1) | 4,489<br>(5.9) | 53,024<br>(100) | 44,972<br>(84.8) | 8,052<br>(15.2) | 68,016<br>(100) | 51,166<br>(75.2) | 16,850<br>(24.8) |
| **Variable (N, % of target outcome)** |  |  |  |  |  |  |  |  |  |  |  |  |
| **Sex** |  |  |  |  |  |  |  |  |  |  |  |  |
| **Female** | 11,255<br>(16) | 10,587<br>(16) | 668<br>(10) | 11,425<br>(15) | 11,255<br>(16) | 170<br>(3.8) | 8,378<br>(16) | 7,043<br>(16) | 1,335<br>(17) | 10,744<br>(16) | 5,215<br>(10) | 5,529<br>(33) |
| **Male** | 60,447<br>(84) | 54,540<br>(84) | 5,907<br>(90) | 64,766<br>(85) | 60,447<br>(84) | 4,319<br>(96) | 44,640<br>(84) | 37,923<br>(84) | 6,717<br>(83) | 57,272<br>(84) | 45,951<br>(90) | 11,321<br>(67) |
| **Race/Ethnicity** |  |  |  |  |  |  |  |  |  |  |  |  |
| **American Indian/Alaska Native, non-Hispanic** | 753<br>(1.1) | 712<br>(1.1) | 41<br>(0.6) | 897<br>(1.2) | 753<br>(1.1) | 144<br>(3.2) | 612<br>(1.2) | 544<br>(1.2) | 68<br>(0.8) | 717 (1.1) | 510<br>(1.0) | 207<br>(1.2) |
| **Asian/Pacific Islander, non-Hispanic** | 806<br>(1.1) | 768<br>(1.2) | 38<br>(0.6) | 868<br>(1.1) | 806<br>(1.1) | 62<br>(1.4) | 601<br>(1.1) | 513<br>(1.1) | 88<br>(1.1) | 773 (1.1) | 526<br>(1.0) | 247<br>(1.5) |
| **Black or African American, non-Hispanic** | 43,357<br>(60) | 38,976<br>(60) | 4,381<br>(67) | 44,546<br>(58) | 43,357<br>(60) | 1,189<br>(26) | 31,293<br>(59) | 25,925<br>(58) | 5,368<br>(67) | 41,109<br>(60) | 33,639<br>(66) | 7,470<br>(44) |
| **Hispanic** | 10,388<br>(14) | 8,745<br>(13) | 1,643<br>(25) | 11,182<br>(15) | 10,388<br>(14) | 794<br>(18) | 8,116<br>(15) | 6,898<br>(15) | 1,218<br>(15) | 9,977<br>(15) | 8,098<br>(16) | 1,879<br>(11) |
| **Two or more races, non-Hispanic** | 632<br>(0.9) | 585<br>(0.9) | 47<br>(0.7) | 718<br>(0.9) | 632<br>(0.9) | 86<br>(1.9) | 460<br>(0.9) | 388<br>(0.9) | 72<br>(0.9) | 605 (0.9) | 434<br>(0.8) | 171<br>(1.0) |
| **White, non-Hispanic** | 15,457<br>(22) | 15,049<br>(23) | 408<br>(6.2) | 17,654<br>(23) | 15,457<br>(22) | 2,197<br>(49) | 11,687<br>(22) | 10,476<br>(23) | 1,211<br>(15) | 14,589<br>(21) | 7,772<br>(15) | 6,817<br>(40) |
| **Age Bins** |  |  |  |  |  |  |  |  |  |  |  |  |
| **Age 15-24** | 19,403<br>(27) | 17,012<br>(26) | 2,391<br>(36) | 20,052<br>(26) | 19,403<br>(27) | 649<br>(14) | 14,421<br>(27) | 11,642<br>(26) | 2,779<br>(35) | 18,454<br>(27) | 15,811<br>(31) | 2,643<br>(16) |
| **Age 25-34** | 21,065<br>(29) | 18,981<br>(29) | 2,084<br>(32) | 22,334<br>(29) | 21,065<br>(29) | 1,269<br>(28) | 15,523<br>(29) | 13,106<br>(29) | 2,417<br>(30) | 20,011<br>(29) | 16,324<br>(32) | 3,687<br>(22) |
| **Age 35-44** | 11,759<br>(16) | 10,907<br>(17) | 852<br>(13) | 12,758<br>(17) | 11,759<br>(16) | 999<br>(22) | 8,595<br>(16) | 7,510<br>(17) | 1,085<br>(13) | 11,136<br>(16) | 8,128<br>(16) | 3,008<br>(18) |
| **Age 45-54** | 6,446<br>(9.0) | 6,088<br>(9.3) | 358<br>(5.4) | 7,074<br>(9.3) | 6,446<br>(9.0) | 628<br>(14) | 4,811<br>(9.1) | 4,303<br>(9.6) | 508<br>(6.3) | 6,097<br>(9.0) | 3,716<br>(7.3) | 2,381<br>(14) |
| **Age 55-64** | 3,545<br>(4.9) | 3,378<br>(5.2) | 167<br>(2.5) | 3,897<br>(5.1) | 3,545<br>(4.9) | 352<br>(7.8) | 2,591<br>(4.9) | 2,333<br>(5.2) | 258<br>(3.2) | 3,330<br>(4.9) | 1,655<br>(3.2) | 1,675<br>(9.9) |
| **Age 65 Plus** | 2,452<br>(3.4) | 2,364<br>(3.6) | 88<br>(1.3) | 2,601<br>(3.4) | 2,452<br>(3.4) | 149<br>(3.3) | 1,806<br>(3.4) | 1,612<br>(3.6) | 194<br>(2.4) | 2,309<br>(3.4) | 687<br>(1.3) | 1,622<br>(9.6) |
| **Unknown** | 6,327<br>(8.8) | 5,771<br>(8.9) | 556<br>(8.5) | 6,766<br>(8.9) | 6,327<br>(8.8) | 439<br>(9.8) | 4,773<br>(9.0) | 4,085<br>(9.1) | 688<br>(8.5) | 5,999<br>(8.8) | 4,486<br>(8.8) | 1,513<br>(9.0) |
| **Intimate Partner**<br>**Violence** |  |  |  |  |  |  |  |  |  |  |  |  |
| **No, Not**<br>**Available,**<br>**Unknown** | 64,071<br>(89) | 57,642<br>(88) | 6,429<br>(98) | 68,095<br>(89) | 64,071<br>(89) | 4,024<br>(90) | 47,096<br>(89) | 39,667<br>(88) | 7,429<br>(92) | 60,535<br>(89) | 48,145<br>(94) | 12,390<br>(74) |
| **Yes** | 7,637<br>(11) | 7,491<br>(12) | 146<br>(2.2) | 8,102<br>(11) | 7,637<br>(11) | 465<br>(10) | 5,928<br>(11) | 5,305<br>(12) | 623<br>(7.7) | 7,481<br>(11) | 3,021<br>(5.9) | 4,460<br>(26) |
| **Mental Health**<br>**Problem** |  |  |  |  |  |  |  |  |  |  |  |  |
| **No, Not**<br>**Available,**<br>**Unknown** | 69,794<br>(97) | 63,323<br>(97) | 6,471<br>(98) | 73,436<br>(96) | 69,794<br>(97) | 3,642<br>(81) | 51,544<br>(97) | 43,633<br>(97) | 7,911<br>(98) | 66,149<br>(97) | 50,059<br>(98) | 16,090<br>(95) |
| **Yes** | 1,914<br>(2.7) | 1,810<br>(2.8) | 104<br>(1.6) | 2,761<br>(3.6) | 1,914<br>(2.7) | 847<br>(19) | 1,480<br>(2.8) | 1,339<br>(3.0) | 141<br>(1.8) | 1,867<br>(2.7) | 1,107<br>(2.2) | 760<br>(4.5) |
| **Alcohol Result** |  |  |  |  |  |  |  |  |  |  |  |  |
| **Not present** | 29,990<br>(42) | 26,479<br>(41) | 3,511<br>(53) | 31,916<br>(42) | 29,990<br>(42) | 1,926<br>(43) | 23,045<br>(43) | 19,419<br>(43) | 3,626<br>(45) | 29,385<br>(43) | 22,307<br>(44) | 7,078<br>(42) |
| **Present** | 16,373<br>(23) | 14,834<br>(23) | 1,539<br>(23) | 17,732<br>(23) | 16,373<br>(23) | 1,359<br>(30) | 12,812<br>(24) | 10,658<br>(24) | 2,154<br>(27) | 16,044<br>(24) | 12,691<br>(25) | 3,353<br>(20) |
| **Unknown / Not**<br>**applicable** | 25,345<br>(35) | 23,820<br>(37) | 1,525<br>(23) | 26,549<br>(35) | 25,345<br>(35) | 1,204<br>(27) | 17,167<br>(32) | 14,895<br>(33) | 2,272<br>(28) | 22,587<br>(33) | 16,168<br>(32) | 6,419<br>(38) |
| **Argument** |  |  |  |  |  |  |  |  |  |  |  |  |
| **No, Not**<br>**Available,**<br>**Unknown** | 54,036<br>(75) | 48,348<br>(74) | 5,688<br>(87) | 57,841<br>(76) | 54,036<br>(75) | 3,805<br>(85) | 39,380<br>(74) | 33,594<br>(75) | 5,786<br>(72) | 50,790<br>(75) | 38,887<br>(76) | 11,903<br>(71) |
| **Yes** | 17,672<br>(25) | 16,785<br>(26) | 887<br>(13) | 18,356<br>(24) | 17,672<br>(25) | 684<br>(15) | 13,644<br>(26) | 11,378<br>(25) | 2,266<br>(28) | 17,226<br>(25) | 12,279<br>(24) | 4,947<br>(29) |

Notes: Outcomes by NVDRS characteristics are displayed in counts and percentages are in parentheses.
Abbreviations for target outcome are: Tot corresponds to total cases, Pos. corresponds to positive class cases, and neg. Corresponds to negative class cases.

Finally, the analysis simulated omitting coroner report text from the training data. Coroner reports may contain extraneous text such as toxicology reports that may be noisy in the context of prediction focused on criminal justice outcomes. Further, compact LLMs have limited token lengths which constrain the number of words in an input narrative and the combination of coroner and homicide reports can exceed the token length in some LLM applications. Because our outcomes are law enforcement focused, the analysis simulated omission of potentially extraneous narrative information.

The analysis began by pre-processing the coroner and police narrative by removing special characters including numbers, punctuation and capitalization as is standard. Police and coroner report narratives were combined into a single field in order to use information available in both narratives (with the exception of the law enforcement narrative-only simulation).

Next, the analysis turned to creating simulated data. First, a test set on which the model outputs were to be evaluated was randomly selected. The test set consisted of a random sample of 30 percent of each outcome’s records, which was then held out from any selection into the training data.

To vary the amounts of training data, the analysis used different training data record counts, each with a different amount of training data. These splits ranged from a minimum of 100 cases, increasing in increments to 200, 500, 1,000, 1,500, and up to 2,000 cases. Each split was randomly sampled from the full dataset specified for each outcome, so that each training split maintained a proportion of positive and negative cases that approximates the true proportion. The prior sample was included in the next iteration to isolate the impact of adding additional training data, not adding different training data. For instance, to obtain 500 cases, first, the prior 200 cases were preserved and an additional 300 were sampled to comprise 500 cases.

To simulate the impacts of language replacement and law enforcement-only text, the analysis followed the procedure process outlined above to randomly select training data in the same 100, 200, 500, 1000, 1500, 2000 increments.

In the second configuration of training data, the composition of positive class cases was altered from the true proportion in the training data. Instead of randomly sampling cases, the proportion of positive class cases was increased in the training data by adding additional positive class cases to the negative class cases. The positive class cases were incrementally increased until they comprise 10, 20, 30, 40, and up to 50 percent of the training data starting from a baseline of 1,000 cases as lower amounts of training data were not performant in this application. For instance, to obtain training data composed of 10 percent positive class cases for legal intervention homicide, the process started with randomly sampled training data with 1,000 records, of which 54 were legal intervention homicides and 940 were not. To the 940 negative class cases, 59 additional positive class cases were added so that the total number of positive class cases was 113 (54+59) and the total was 1,059 cases, of which approximately 10 percent (113 / 1,059) were legal intervention homicides.

For each of the configurations described above, distilBERT, a LLM with fewer parameters but comparable accuracy to large scale LLMs, was used.^45^ Compact LLMs in this context were selected to better allow for simulated iteration with fewer computational needs and because data protections do not data cloud storage and computing application. The distilBERT models were fine-tuned on training data to select model parameters. Parameters were selected in initial fine-turning using two outcomes (legal intervention and drive-by). Because model parameters in each fine-tuned model were identical, these parameters were applied to each training data configuration (see Appendix Table 2). Because our target outcomes are imbalanced, we add a weighted trainer to account for class imbalance.

**Table 2:** Narrative Descriptive Statistics, Characteristics by Outcome.

|  | Driveby |  |  | Legal Intervention |  |  | Number Nonfatally Shot |  |  | Victim Injured at Home |  |  |
| --- | --- | --- | --- | --- | --- | --- | --- | --- | --- | --- | --- | --- |
| Variable | Tot | Neg | Pos | Tot | Neg | Pos | Tot | Neg | Pos | Tot | Neg | Pos |
| **Panel A:**<br>**Overall** |  |  |  |  |  |  |  |  |  |  |  |  |
| Circumstances Known, N () | 54,354 (76) | 47,779 (73) | 6,575 (100) | 58,745 (77) | 54,354 (76) | 4,391 (98) | 41,794 (79) | 34,852 (77) | 6,942 (86) | 52,852 (78) | 38,879 (76) | 13,973 (83) |
| Words Law Enforcement Narrative, Median (IQR) | 80 (37, 137) | 78 (35, 136) | 93 (53, 148) | 81 (37, 141) | 80 (37, 137) | 115 (44, 206) | 83 (41, 141) | 78 (39, 134) | 113 (65, 173) | 83 (40, 141) | 80 (39, 132) | 94 (45, 168) |
| Words CME Narrative, Median (IQR) | 89 (55, 138) | 88 (54, 137) | 100 (64, 147) | 90 (56, 141) | 89 (55, 138) | 120 (78, 182) | 88 (56, 137) | 84 (53, 132) | 108 (72, 163) | 91 (58, 140) | 89 (56, 134) | 100 (62, 161) |
| **Panel B:**<br>**Black** |  |  |  |  |  |  |  |  |  |  |  |  |
| Circumstances Known, N () | 30,774 (71) | 26,393 (68) | 4,381 (100) | 31,930 (72) | 30,774 (71) | 1,156 (97) | 23,041 (74) | 18,562 (72) | 4,479 (83) | 29,875 (73) | 24,054 (72) | 5,821 (78) |
| Words Law Enforcement Narrative, Median (IQR) | 77 (38, 125) | 74 (36, 121) | 98 (59, 152) | 77 (38, 126) | 77 (38, 125) | 98 (41, 169) | 80 (42, 126) | 74 (39, 118) | 109 (67, 160) | 80 (41, 127) | 79 (41, 125) | 82 (43, 137) |
| Words CME Narrative, Median (IQR) | 85 (55, 126) | 83 (53, 123) | 105 (72, 151) | 86 (55, 127) | 85 (55, 126) | 105 (73, 150) | 84 (56, 125) | 80 (53, 119) | 106 (72, 155) | 87 (57, 128) | 87 (57, 126) | 89 (58, 137) |
| **Panel C:**<br>**Hispanic** |  |  |  |  |  |  |  |  |  |  |  |  |
| Circumstances Known, N () | 8,698 (84) | 7,055 (81) | 1,643 (100) | 9,487 (85) | 8,698 (84) | 789 (99) | 7,107 (88) | 5,980 (87) | 1,127 (93) | 8,521 (85) | 6,906 (85) | 1,615 (86) |
| Words Law Enforcement Narrative, Median (IQR) | 67 (29, 136) | 66 (24, 139) | 72 (40, 129) | 69 (28, 144) | 67 (29, 136) | 132 (13, 247) | 69 (34, 142) | 65 (32, 132) | 106 (52, 186) | 69 (31, 139) | 66 (31, 130) | 89 (32, 186) |
| Words CME Narrative, Median (IQR) | 87 (45, 146) | 88 (48, 149) | 79 (31, 134) | 91 (47, 151) | 87 (45, 146) | 139 (90, 206) | 79 (41, 141) | 75 (39, 134) | 106 (60, 170) | 88 (47, 147) | 83 (44, 141) | 111 (66, 180) |
| **Panel D:**<br>**White** |  |  |  |  |  |  |  |  |  |  |  |  |
| Circumstances Known, N (%) | 12,860 (83) | 12,452 (83) | 408 (100) | 15,003 (85) | 12,860 (83) | 2,143 (98) | 10,052 (86) | 8,952 (85) | 1,100 (91) | 12,496 (86) | 6,537 (84) | 5,959 (87) |
| Words Law Enforcement Narrative, Median (IQR) | 99 (42, 182) | 99 (41, 181) | 112 (65, 186) | 102 (43, 186) | 99 (42, 182) | 119 (53, 212) | 105 (48, 186) | 101 (47, 181) | 134 (76, 232) | 104 (47, 187) | 97 (43, 172) | 114 (52, 205) |
| Words CME Narrative, Median (IQR) | 103 (61, 165) | 102 (61, 165) | 107 (72, 149) | 105 (63, 167) | 103 (61, 165) | 122 (77, 184) | 104 (65, 166) | 103 (63, 163) | 118 (74, 191) | 106 (65, 168) | 101 (63, 156) | 112 (68, 183) |
| **Panel E: Female** |  |  |  |  |  |  |  |  |  |  |  |  |
| Circumstances Known, N (%) | 9,404 (84) | 8,736 (83) | 668 (100) | 9,567 (84) | 9,404 (84) | 163 (96) | 7,242 (86) | 6,065 (86) | 1,177 (88) | 9,169 (85) | 4,271 (82) | 4,898 (89) |
| Words Law Enforcement Narrative, Median (IQR) | 104 (48, 184) | 104 (47, 186) | 109 (60, 158) | 104 (48, 184) | 104 (48, 184) | 122 (49, 193) | 109 (54, 189) | 106 (51, 186) | 125 (70, 204) | 107 (52, 187) | 97 (47, 165) | 118 (57, 207) |
| Words CME Narrative, Median (IQR) | 111 (68, 181) | 112 (68, 183) | 105 (67, 152) | 111 (68, 181) | 111 (68, 181) | 134 (90, 198) | 113 (71, 181) | 111 (69, 181) | 122 (80, 188) | 113 (71, 184) | 105 (66, 165) | 124 (75, 198) |
| **Panel F: Male** |  |  |  |  |  |  |  |  |  |  |  |  |
| Circumstances Known, N (%) | 44,950 (74) | 39,043 (72) | 5,907 (100) | 49,178 (76) | 44,950 (74) | 4,228 (98) | 34,552 (77) | 28,787 (76) | 5,765 (86) | 43,683 (76) | 34,608 (75) | 9,075 (80) |
| Words Law Enforcement Narrative, Median (IQR) | 76 (35, 130) | 74 (34, 128) | 92 (52, 147) | 78 (36, 134) | 76 (35, 130) | 114 (44, 207) | 79 (40, 133) | 74 (37, 125) | 110 (64, 168) | 79 (39, 133) | 78 (39, 129) | 85 (41, 150) |
| Words CME Narrative, Median (IQR) | 86 (53, 131) | 84 (53, 129) | 100 (64, 146) | 87 (55, 135) | 86 (53, 131) | 120 (78, 181) | 84 (54, 129) | 81 (52, 124) | 106 (70, 158) | 88 (56, 133) | 87 (55, 131) | 92 (58, 145) |

Notes: Outcomes by NVDRS characteristics are displayed in counts and percentages for categorical outcomes including known circumstances. For numeric outcomes, the median is presented along with the interquartile range (25th and 75th percentile) in parentheses. Abbreviations for target outcome are: Tot corresponds to total cases, Pos. corresponds to positive class cases, and neg. Corresponds to negative class cases.

Classification performance was measured using learning curves, which plot performance metrics relative to differing splits of labeled training data to evaluate classifier model performance. Binary classification model metrics including precision and recall in addition to metrics considered useful for imbalanced class problems, including an F1 score, were used. Finally, to analyze classification performance by subgroup, learning curves were created for sex race, and ethnicity subgroups.

## RESULTS

Classification outcomes differed by the proportion of positive to negative cases in each outcome (Table 1). The most rare positive class outcome was a police shooting (5.9%) followed by drive-by shootings (9.2%) and shootings where additional victims were non fatally shot (15%) in the course of the homicide. The most prevalent outcome was whether a victim is shot in their home (25%) relative to another location outside the home. Victims of homicide in the sample tended to be male (84-85%), Black or African American (58-60%), and young, with the most frequent age range between 25-34 years of age (Table 1). Intimate partner violence characterizes over a tenth of homicides overall but within cases where a victim is injured at home, intimate partner violence and preceding arguments occurred in over a quarter of cases (26% and 29%). Legal intervention homicides were most likely associated with mental health problems and alcohol use.

Circumstances were known for almost all cases of legal intervention and drive-by shootings (98 and 100%) but less information was known about the circumstances of homicide where additional victims were shot or when victims were injured at home (Table 2). Circumstances were known in 71% of homicides of Black victims in contrast to 83-84% among Hispanic and non-Hispanic white victims. The median number of words in a narrative for a law enforcement narrative was 81-83 words whereas CME narratives ranged from 88-91 words in length. Legal intervention homicides had the most lengthy narratives (115 for LE and 120 for CME). Narrative length differed by race and sex. Among law enforcement narratives, median length for Black victims was 98 words but 132 for non-Hispanic white victims. Narrative length differed among male and female victims. Female victims had longer narratives for each homicide outcome. Female victims shot at home had a median narrative length of 124 words in contrast to male victims shot at home with a length of 92 words.

Table 3 displays classification performance by F1 score for each model type. Training data of approximately 1,500 cases achieved an F1 score of at least .6 for each outcome, though at 1,000 cases the majority of outcomes were at or exceeding .6. The exception was the number non-fatally shot. Figure 1 plots learning curves by F1 score in Table 3. Replacement language models tended to perform best (Table 3, Figure 1) with the highest F1 score in all save six model interactions. In particular, language replacement models consistently obtained the highest F1 score for legal intervention homicides (Table 3, Figure 1). Omitting coroner/ medical examiner reports performed worse across outcomes. Language replacement models trained on 1,500-2,000 narratives obtained low false negative rates ranging from 1-5% of true cases resulting in a misclassified outcome (Figure 1, Appendix Table 4).

**Table 3:** F1 Scores by Model Outcome, Training Data, and Model Type.

| **Outcome** | **Train N** | **distilBERT** | **distilBERT + LE only** | **distilBERT + language** |
| --- | --- | --- | --- | --- |
| Driveby | 100 | **0.219** | 0.168 | 0.209 |
| Driveby | 200 | **0.232** | 0.148 | 0.232 |
| Driveby | 500 | 0.381 | 0.144 | **0.473** |
| Driveby | 1000 | **0.626** | 0.124 | 0.606 |
| Driveby | 1500 | 0.619 | 0.126 | **0.623** |
| Driveby | 2000 | 0.593 | 0.126 | **0.635** |
| Police Shooting | 100 | 0.231 | 0.105 | **0.305** |
| Police Shooting | 200 | 0.218 | 0.083 | **0.364** |
| Police Shooting | 500 | 0.490 | 0.083 | **0.653** |
| Police Shooting | 1000 | 0.739 | 0.064 | **0.795** |
| Police Shooting | 1500 | 0.771 | 0.056 | **0.856** |
| Police Shooting | 2000 | 0.770 | 0.080 | **0.833** |
| Number Nonfatally Shot | 100 | **0.319** | 0.246 | 0.312 |
| Number Nonfatally Shot | 200 | 0.281 | 0.226 | **0.286** |
| Number Nonfatally Shot | 500 | 0.341 | 0.192 | **0.345** |
| Number Nonfatally Shot | 1000 | 0.352 | 0.220 | **0.413** |
| Number Nonfatally Shot | 1500 | 0.591 | 0.182 | **0.642** |
| Number Nonfatally Shot | 2000 | 0.608 | 0.195 | **0.663** |
| Victim Injured at Home | 100 | 0.547 | 0.222 | **0.574** |
| Victim Injured at Home | 200 | 0.578 | 0.283 | **0.629** |
| Victim Injured at Home | 500 | 0.665 | 0.277 | **0.714** |
| Victim Injured at Home | 1000 | **0.722** | 0.294 | 0.697 |
| Victim Injured at Home | 1500 | 0.737 | 0.280 | **0.749** |
| Victim Injured at Home | 2000 | **0.744** | 0.286 | 0.739 |

Notes: Each cell contains an F1 score produced by each model. Model type is denoted in columns 3-5, where column 3 is the base distilBERT model, column 4 is the distilBERT model trained only on law enforcement narratives and column 5 is the distilBERT model where text replacement for uncommon language in the narratives is replaced for clarity. Cells that are bolded correspond to the best F1 score across the listed models.

**Figure 1:**
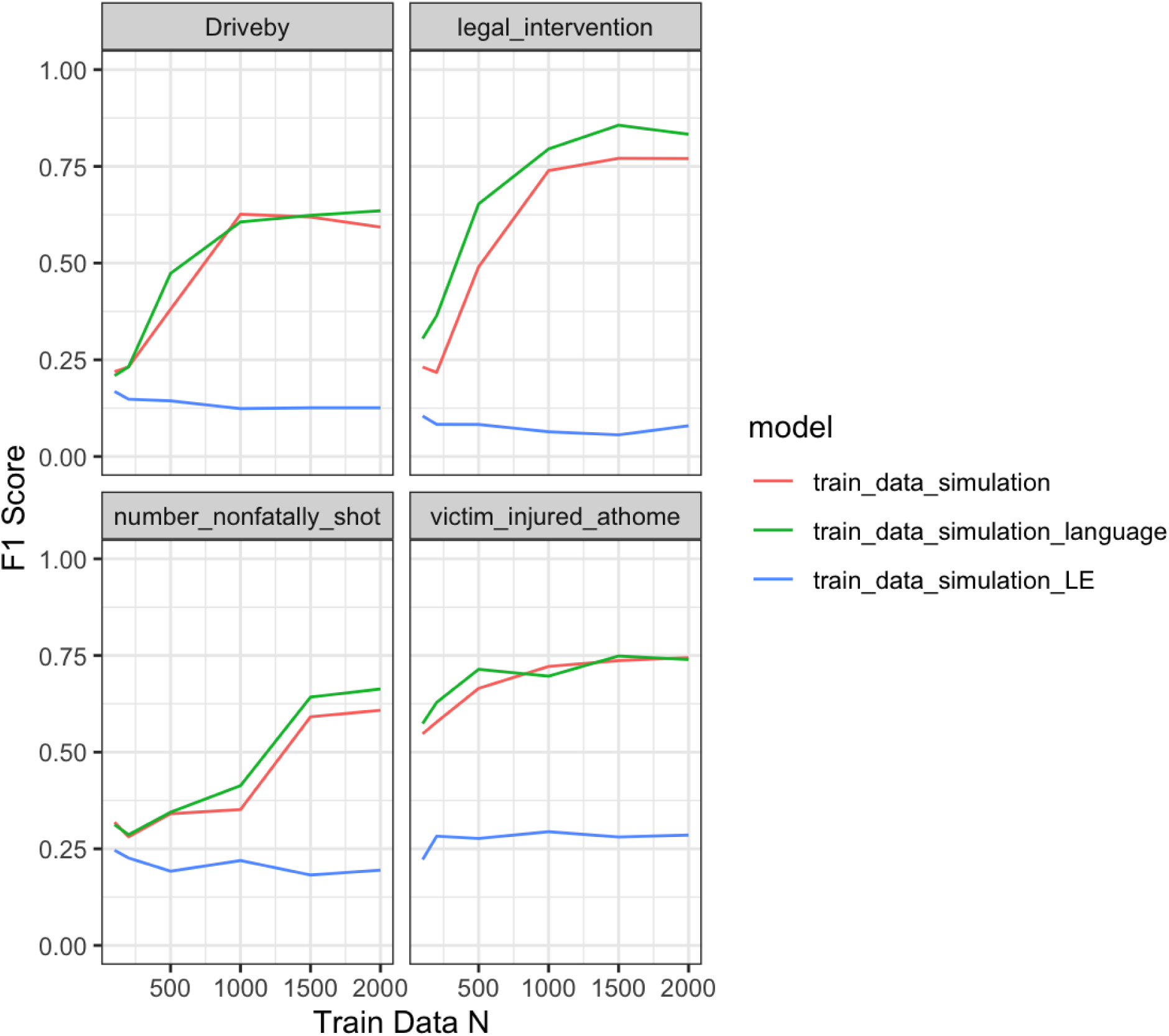
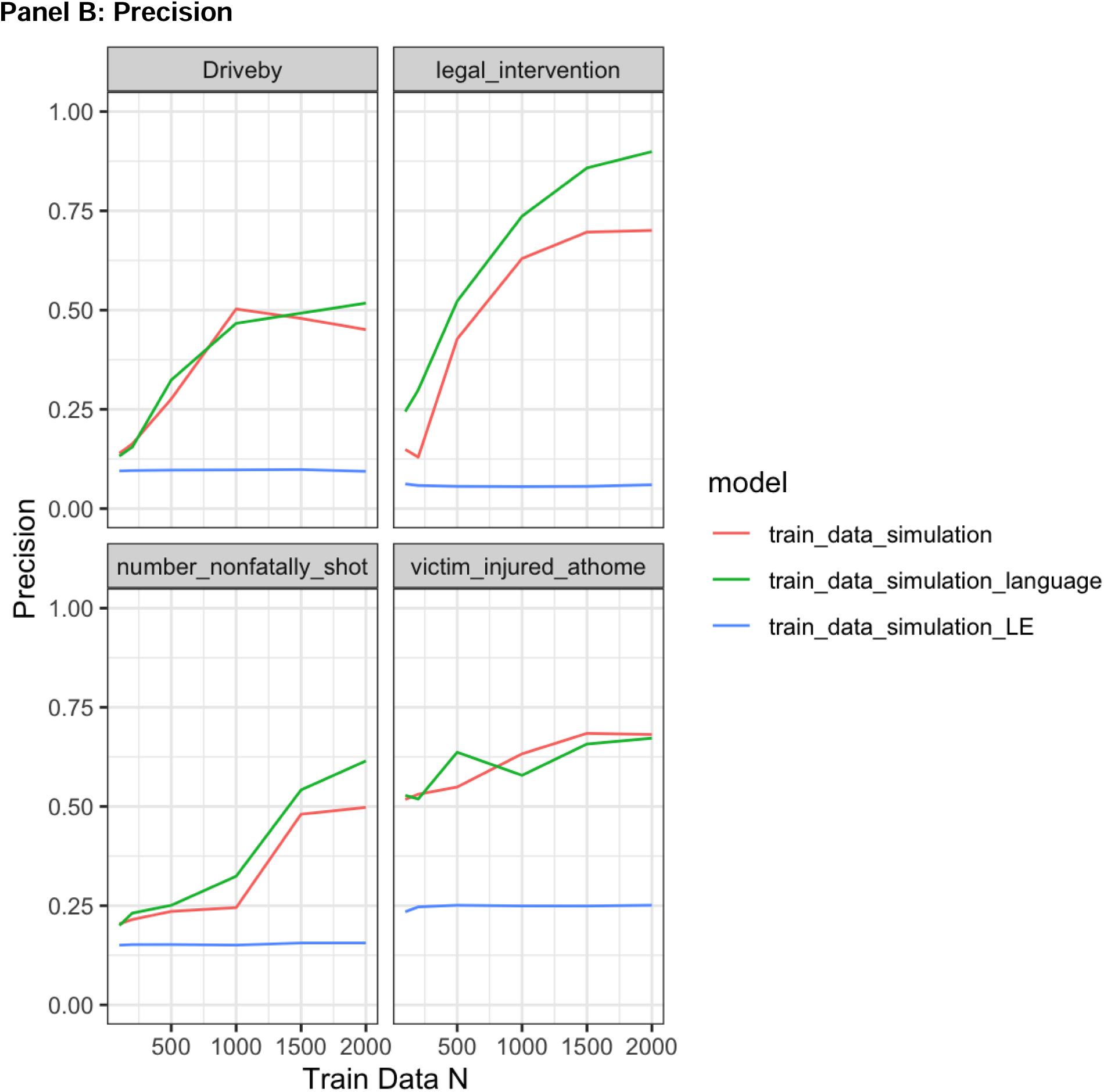
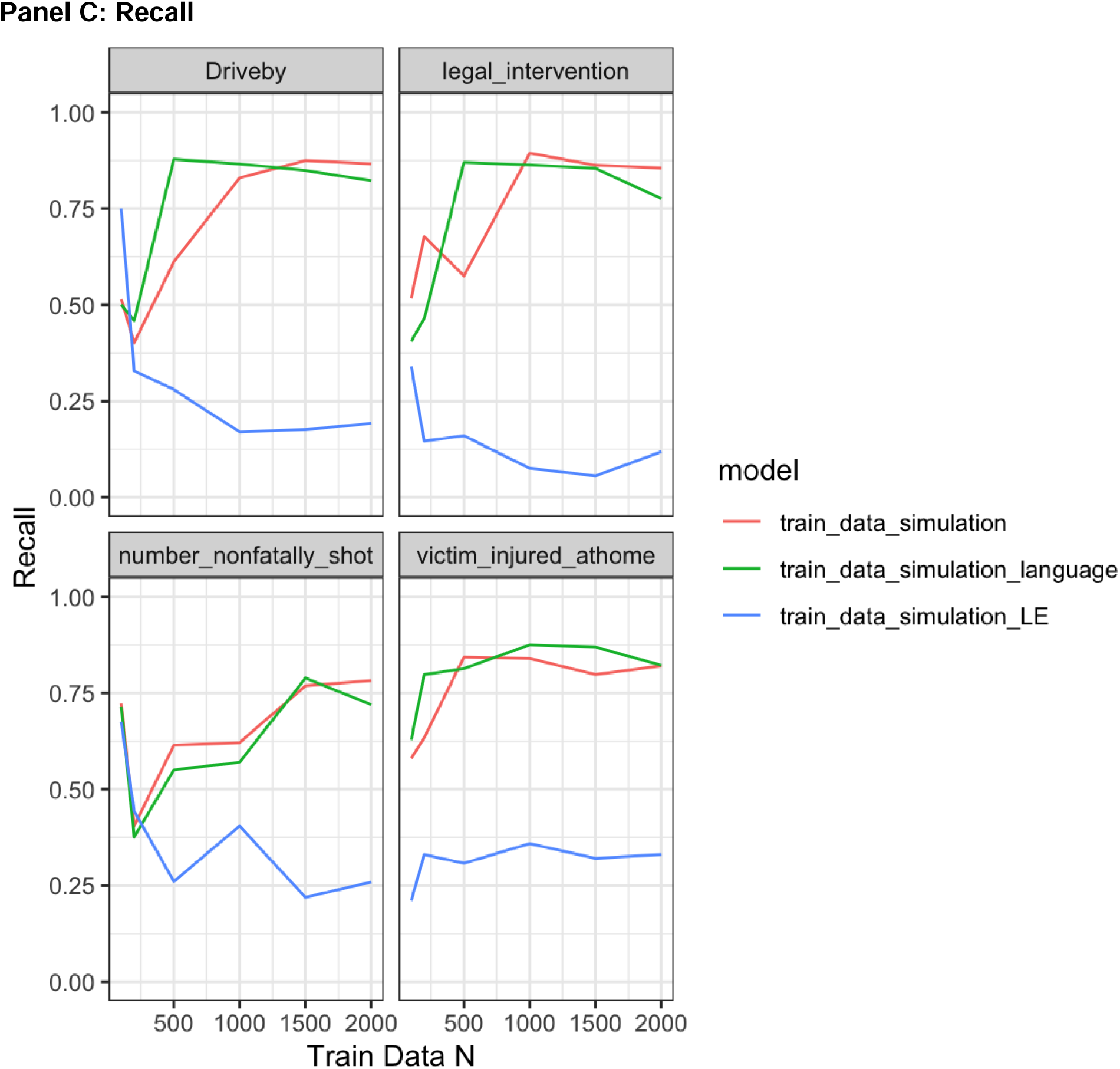
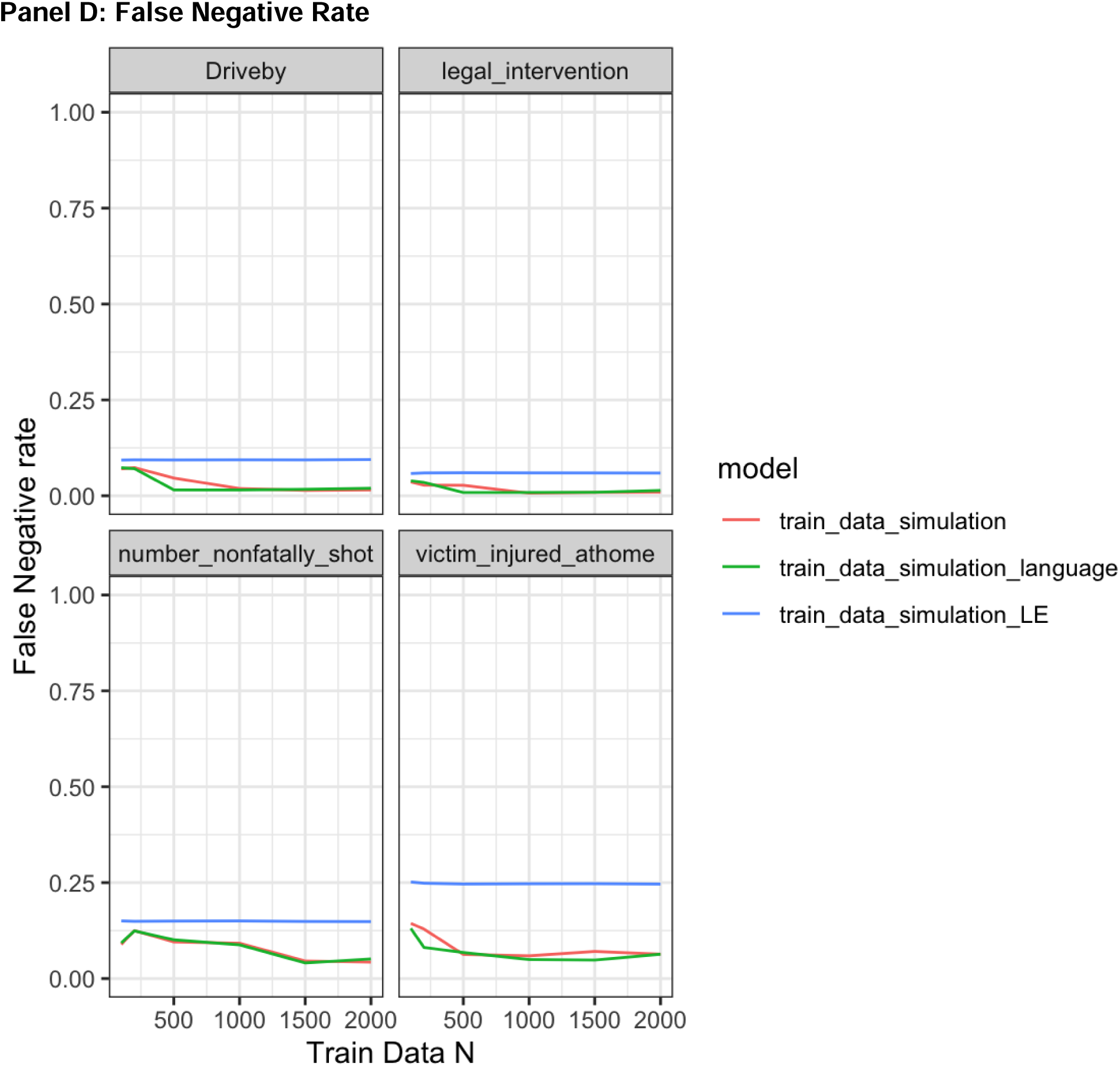
Learning Curve by Outcome, Model Type Panel A: F1 Score.

**Table 4:** Classification Performance for Language Replacement Models by Outcome by Subgroup.

| Category | Train N | F1 - white | F1 - Black | F1 - Hispanic | F1 - Male | F1 - Female |
| --- | --- | --- | --- | --- | --- | --- |
| Driveby | 100 | 0.208 | 0.353 | 0.322 | 0.317 | 0.292 |
| Driveby | 200 | 0.204 | 0.321 | 0.280 | 0.292 | 0.266 |
| Driveby | 500 | 0.237 | 0.390 | 0.346 | 0.356 | 0.302 |
| Driveby | 1000 | 0.321 | 0.447 | 0.380 | 0.416 | 0.401 |
| Driveby | 1500 | 0.536 | 0.675 | 0.607 | 0.651 | 0.600 |
| Driveby | 2000 | 0.554 | 0.704 | 0.608 | 0.672 | 0.622 |
| Legal Intervention | 100 | 0.383 | 0.169 | 0.324 | 0.336 | 0.085 |
| Legal Intervention | 200 | 0.475 | 0.162 | 0.411 | 0.391 | 0.138 |
| Legal Intervention | 500 | 0.746 | 0.473 | 0.722 | 0.678 | 0.343 |
| Legal Intervention | 1000 | 0.812 | 0.723 | 0.816 | 0.817 | 0.495 |
| Legal Intervention | 1500 | 0.852 | 0.830 | 0.893 | 0.870 | 0.632 |
| Legal Intervention | 2000 | 0.833 | 0.793 | 0.873 | 0.842 | 0.672 |
| Number Nonfatally Shot | 100 | 0.075 | 0.248 | 0.238 | 0.226 | 0.127 |
| Number Nonfatally Shot | 200 | 0.073 | 0.290 | 0.268 | 0.255 | 0.131 |
| Number Nonfatally Shot | 500 | 0.239 | 0.478 | 0.604 | 0.479 | 0.431 |
| Number Nonfatally Shot | 1000 | 0.382 | 0.613 | 0.693 | 0.613 | 0.558 |
| Number Nonfatally Shot | 1500 | 0.412 | 0.629 | 0.688 | 0.626 | 0.602 |
| Number Nonfatally Shot | 2000 | 0.425 | 0.640 | 0.712 | 0.639 | 0.610 |
| Victim Injured at Home | 100 | 0.679 | 0.488 | 0.543 | 0.505 | 0.724 |
| Victim Injured at Home | 200 | 0.733 | 0.552 | 0.605 | 0.566 | 0.783 |
| Victim Injured at Home | 500 | 0.785 | 0.660 | 0.680 | 0.665 | 0.824 |
| Victim Injured at Home | 1000 | 0.784 | 0.637 | 0.649 | 0.642 | 0.826 |
| Victim Injured at Home | 1500 | 0.821 | 0.709 | 0.663 | 0.703 | 0.851 |
| Victim Injured at Home | 2000 | 0.805 | 0.696 | 0.683 | 0.699 | 0.828 |

Oversampling positive class cases was negligibly helpful in improving F1 scores (Figure 2). For instance, oversampling for legal intervention homicide to be composed of 20 percent positive class cases resulted in the addition of 580 positive class cases added to training data and an F1 score of .795 (Appendix Table 4; Figure 2). Relative to adding 500 randomly sampled cases which would result in an F1 score of .771 (Appendix Table 4), the gain from oversampling was 0.024 (.795 - .771) and therefore modest.

**Figure 2:**
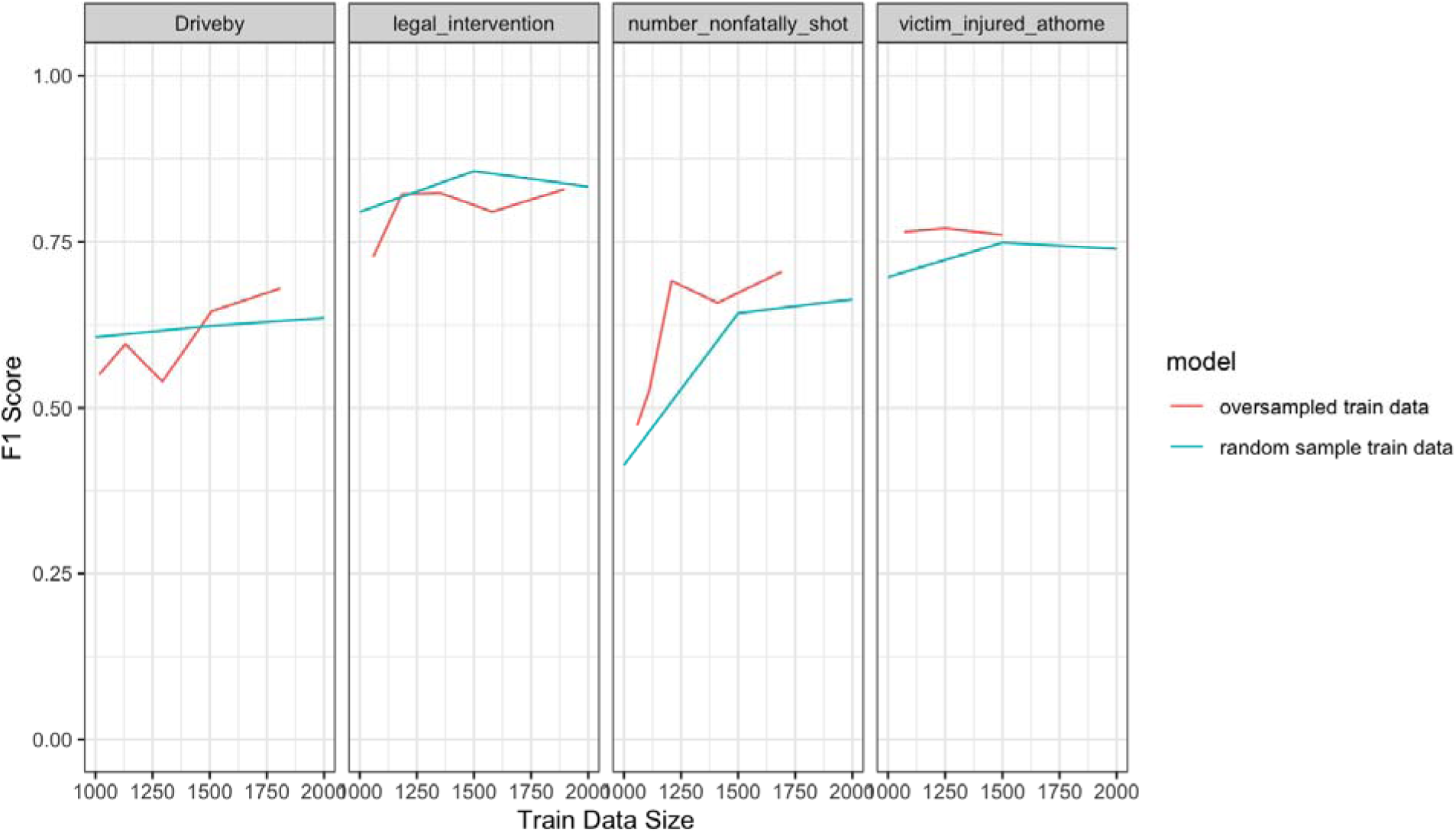
F1 Learning Curve for Oversampled Positive Class Cases vs. Baseline Language Replacement Model. Notes: F1 scores are plotted for distilBERT models fit with language replacement for both randomly sampled training data and oversampled training data. Oversampled training data corresponds to an increment of a 10 percent increase in the proportion of positive class cases included in training data. Exact training data set counts are in Appendix Table 4. Random train data is plotted at n=1,000, 1,500 and 2,000 randomly sampled training data records for reference.

Figure 3 plots F1 scores of distilBERT language replacement models as these models tended to perform best overall and may capture linguistic differences most accurately across subgroups. Predictions differ by race/ethnicity and sex across models. Legal intervention homicide victims who were white or Hispanic were most often correctly classified as such, and Black victims were least likely to be correctly classified (Figure 3, panel A). The prediction difference is substantial for legal intervention victims with lower amounts of training data, though the gap persisted with higher volumes of training data. White victims shot at home were most often correctly predicted while Black and Hispanic victims were least likely. Female victims were less likely to be correctly predicted than male victims in all instances save if they were shot at home. Among models with at least 1,500 records of training data, F1-score disparities ranged from 0.2 to 0.25 by race and ethnicity, and between male and female victims with differences ranging from 0.12 to 0.2 (Table 4).

**Figure 3:**
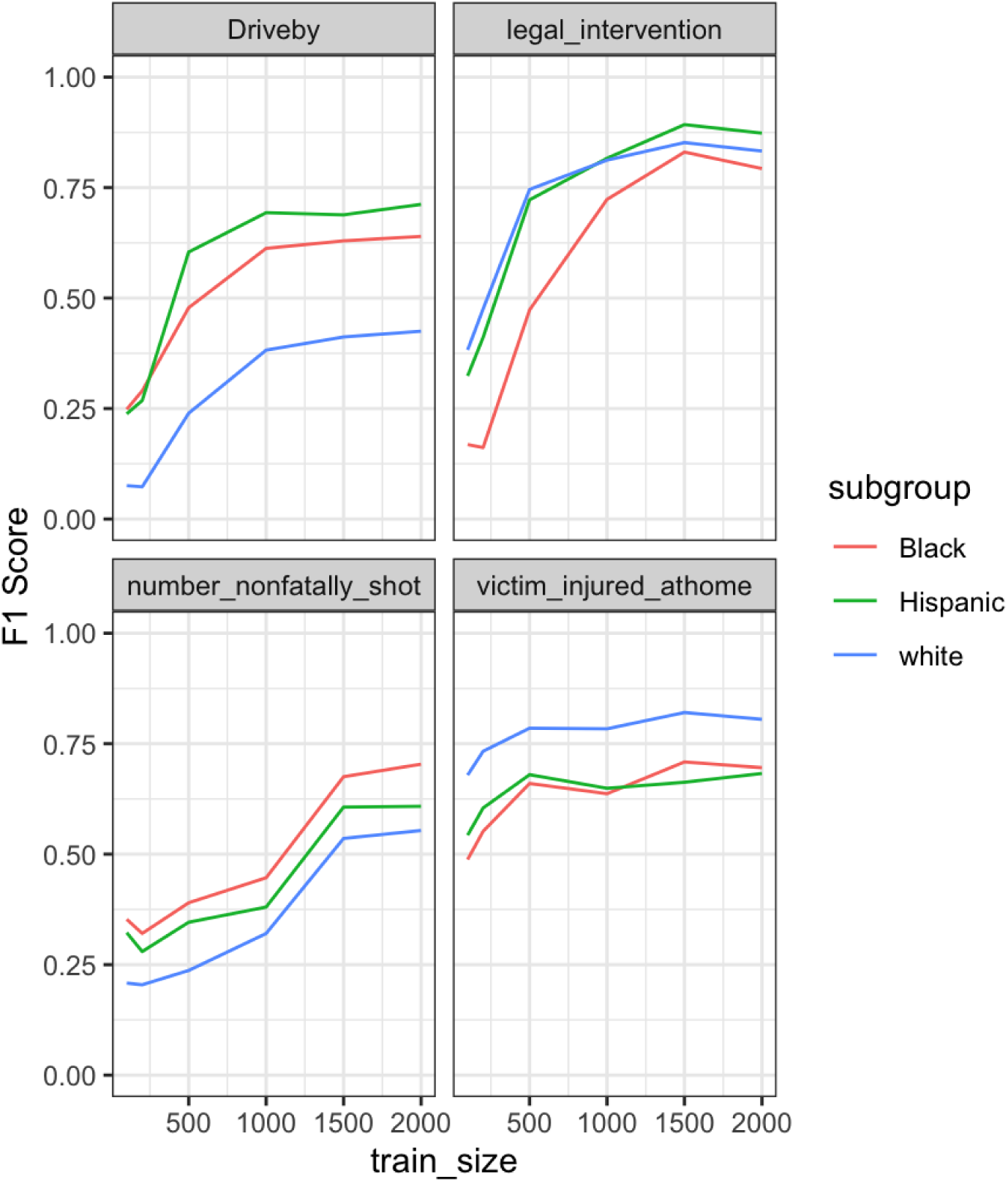
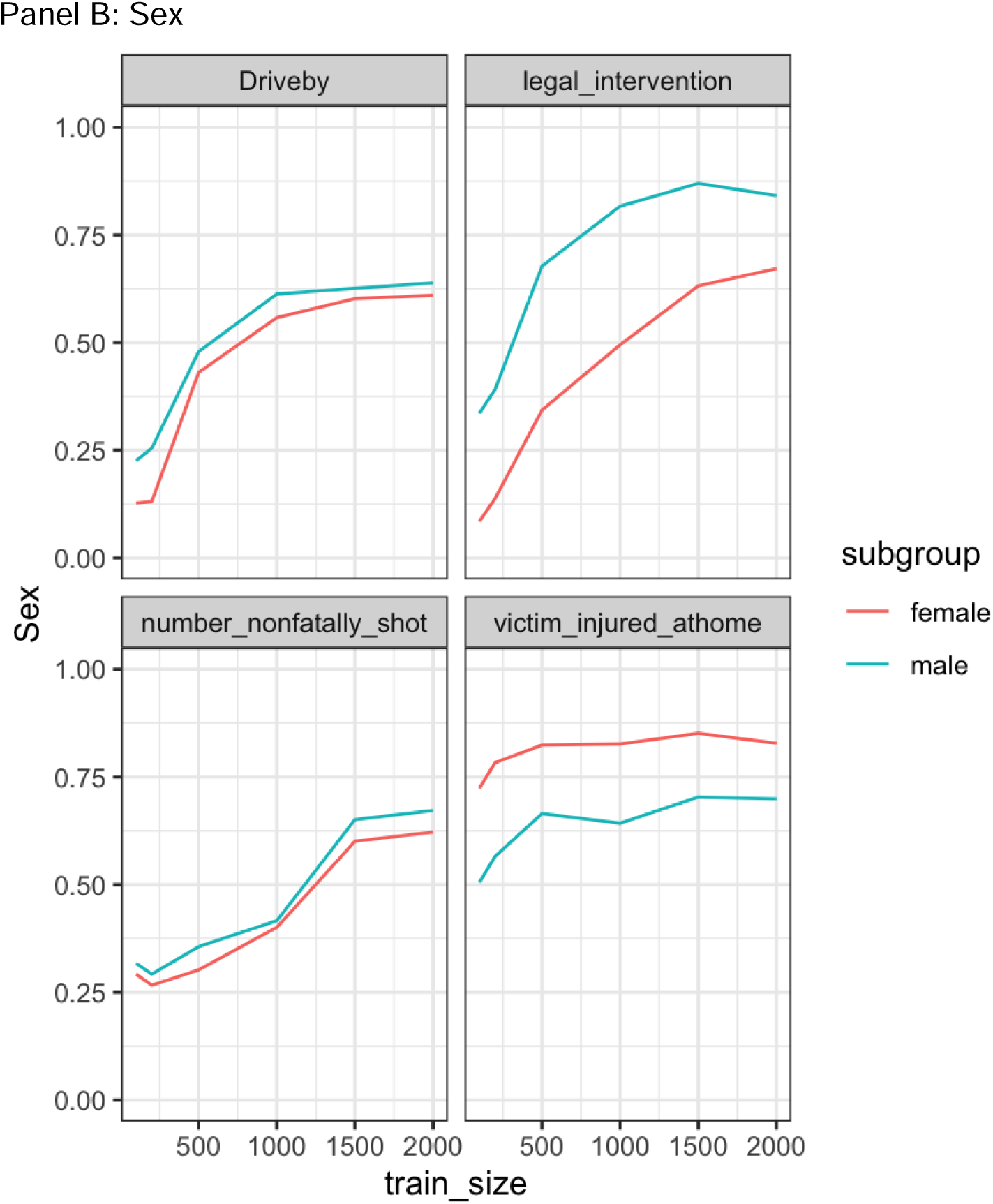
F1 Learning Curves for distilBERT+language models by Subgroup.

## DISCUSSION

This analysis simulated the NLP model-fitting process to demonstrate how different training and pre-processing decisions impact model performance in NLP applications of text classification of violent death homicide and coroner narratives. Fine tuning compact LLMs on NVDRS text requires approximately 1,000-1,500 training data records to achieve an F1 score of at least .6. Results suggested that compact LLMs are less useful in few shot learning applications with limited training data. Oversampling the positive class cases in training data does not increase prediction accuracy substantially over randomly sampled training data.

Predictions differed by race, ethnicity and sex. Differential prediction by subgroup is not explainable by outcome frequency or narrative length alone. For instance, white victims of police shootings are less prevalent than Black victims in the sample but are more often classified correctly. Similarly, female victims have longer median narratives for all outcomes, but are less likely to be correctly classified. Further research should characterize sources of differential prediction, whether input narratives or exacerbation by NLP classifier, and examine fairness aware models particularly if the prediction is used for decision-making or resource allocation in public health settings.

These findings may inform a range of researchers and practitioners in the health informatics and public health. Compact LLMs with simple text changes can effectively predict rare NVDRS outcomes is of use to researchers considering the use of supervised machine learning to expand what is known about violent deaths beyond existing coded fields. For researchers seeking annotated training data, random sampling and labeling a sufficient number of cases (approximately 1,000) combined with a weighting layer is an effective strategy. Further, manual annotation requirements for NVDRS applications to rare events do not require prohibitive amounts of training data which can require substantial costs. For instance, if labeling a narrative requires approximately 2-5 minutes of annotator time, it would require approximately 33-83 hours of annotation time for one annotator. While it is likely that future access to privacy-compliant sophisticated LLMs will be more accessible to researchers working with sensitive data, in the interim, this analysis provides useful baselines for researchers considering similar undertakings. Finally, this approach may assist state level violent death reporting systems in the lengthy process of abstraction in NVDRS, where manual abstractor annotation of violent death narratives result in long delays to data access.

This research is subject to several limitations. First, results from a compact LLM may not fully generalize to new LLMs with additional sophistication or to different language contexts beyond NVDRS. Label noise from NVDRS annotators may mean that results understate the performance of compact LLMs, which is consistent with police shootings tending to be the outcome type that is most accurately predicted. The potential for differential prediction by subgroup raises concerns about fairness and equity in model performance. Further investigations into the sources of this differential prediction are needed to ensure that NLP applications do not exacerbate existing disparities.

## Conclusion

This study conducted a comprehensive analysis of the model-fitting process for supervised binary classification of infrequent violent death outcomes using natural language processing (NLP) techniques on National Violent Death Reporting System (NVDRS) narrative data. Through simulations, this study examined the impacts of pre-processing, the quantity and composition of labeled training data on model performance, as well as by race, ethnicity, and sex subgroups.

## Data Availability

All data produced in the study are available for permissioned researchers by applying for restricted access National Violent Death Reporting System Data.

https://www.cdc.gov/nvdrs/about/nvdrs-data-access.html

## Acknowledgments

I thank Matthew Miller and Deb Azrael for comments on a prior draft of this article. I thank Daniel Bowen and Stephen Sumner for valuable discussion in development of this article.

## Competing Interest

None

Funding and all other required statements

This work was funded by APHA AWARD # 2023-0011.

## Notes

### Competing Interest Statement

The authors have declared no competing interest.

### Funding Statement

This study was funded by CDC award NU38OT000294 and the American Public Health Association (APHA) AWARD # 2023-0011

