## Supplement for "Assessing Supervised Natural Language Processing (NLP) Classification of Violent Death Narratives: Development and Assessment of a Compact Large Language Model (LLM) Approach"

Appendix Table 1: Language Replacement words and replacement value

| Replaced Words | Value |
|---|---|
| Gsw, gsws, gun shot wound | Gunshot wound |
| Vic, V, V's, Vic's | Victim |
| S, S's | Suspect |
| Law enforcement, LE, LEO, officer, police officer, officers, cop, cops, sheriff, sheriffs, detective, trooper, troopers, marshall, marshals , deputy, deputies | Police |

Appendix Table 2: Model Parameters

| Parameter | Value |
|---|---|
| Learning rate | 0.0005 |
| Batch size | 60 |
| Number of epochs | 5 |
| Weight decay | 0.01 |

Appendix Table 3: Summary of Simulations

| Simulation | Type | Training Data | Training Data N |
|---|---|---|---|
| 1 - distilBERT | Model fitting | Random sample | 100, 200, 500, 1000, 1500, 2000 |
| 2 - distilBERT + oversample | Pre-processing | Oversampled positive class outcome | 1000, 10%, 20%, 30% , 40%, 50% |
| 3 - Distilbert + Language | Pre-processing | Random Sample; Language replacement | 100, 200, 500, 1000, 1500, 2000 |
| 4 - distilBERT + LE only | Pre-processing | Random Sample; Law enforcement narrative only | 100, 200, 500, 1000, 1500, 2000 |

**Appendix Table 4: Classification Performance by Oversampled Proportion of Positive Class Case**

| Outcome | Train Proportion | Train N | F1 | Precision | Recall |
|---|---|---|---|---|---|
| Driveby | Random Sample | 1000 | 0.626 | 0.503 | 0.830 |
| Driveby | 10 percent | 1016 | 0.550 | 0.419 | 0.802 |
| Driveby | 20 percent | 1131 | 0.596 | 0.451 | 0.877 |
| Driveby | 30 percent | 1293 | 0.539 | 0.383 | 0.913 |
| Driveby | 40 percent | 1508 | 0.645 | 0.513 | 0.869 |
| Driveby | 50 percent | 1810 | **0.680** | 0.598 | 0.788 |
| Police Shooting | Random Sample | 1000 | 0.739 | 0.630 | 0.894 |
| Police Shooting | 10 percent | 1059 | 0.727 | 0.633 | 0.854 |
| Police Shooting | 20 percent | 1185 | 0.822 | 0.745 | 0.916 |
| Police Shooting | 30 percent | 1354 | 0.823 | 0.741 | 0.926 |
| Police Shooting | 40 percent | 1580 | 0.795 | 0.682 | 0.954 |
| Police Shooting | 50 percent | 1896 | **0.829** | 0.734 | 0.953 |
| Number Nonfatally Shot | Random Sample | 1000 | 0.352 | 0.245 | 0.621 |
| Number Nonfatally Shot | 10 percent | 1111 | 0.526 | 0.506 | 0.547 |
| Number Nonfatally Shot | 20 percent | 1058 | 0.473 | 0.357 | 0.701 |
| Number Nonfatally Shot | 30 percent | 1209 | 0.691 | 0.616 | 0.786 |
| Number Nonfatally Shot | 40 percent | 1410 | 0.658 | 0.530 | 0.868 |
| Number Nonfatally Shot | 50 percent | 1692 | **0.705** | 0.597 | 0.860 |
| Victim Injured at Home | Random Sample | 1000 | 0.722 | 0.633 | 0.840 |
| Victim Injured at Home | 30 percent | 1071 | **0.765** | 0.734 | 0.799 |
| Victim Injured at Home | 40 percent | 1250 | 0.770 | 0.747 | 0.795 |
| Victim Injured at Home | 50 percent | 1500 | 0.761 | 0.679 | 0.864 |

Notes: Each numeric cell contains performance (F1, precision, recall) for each model outcome by the proportion of positive class cases that were included in the training data. In Column 2, the proportion of the training data that was a positive class case is listed, with the exception being “random sample” which denotes training data that is randomly sampled up to n=1000 cases and is presented for reference. The train proportion increases the proportion of positive class cases beyond the random sample. The exception is the outcome Victim Injured at Home which omits the train proportion at 10 and 20 percent because the randomly sampled proportion exceeds 20 percent.

**Appendix Table 5: Classification Metrics for Language Replacement Models**

| Category | Train N | F1 | Precision | Recall | FNR |
|---|---|---|---|---|---|
| Driveby | 100 | 0.21 | 0.13 | 0.5 | 0.07 |
| Driveby | 200 | 0.23 | 0.15 | 0.46 | 0.07 |
| Driveby | 500 | 0.47 | 0.32 | 0.88 | 0.02 |
| Driveby | 1000 | 0.61 | 0.47 | 0.87 | 0.02 |
| Driveby | 1500 | 0.62 | 0.49 | 0.85 | 0.02 |
| Driveby | 2000 | 0.64 | 0.52 | 0.82 | 0.02 |
| Legal Intervention | 100 | 0.3 | 0.24 | 0.41 | 0.04 |
| Legal Intervention | 200 | 0.36 | 0.3 | 0.46 | 0.03 |
| Legal Intervention | 500 | 0.65 | 0.52 | 0.87 | 0.01 |
| Legal Intervention | 1000 | 0.79 | 0.74 | 0.86 | 0.01 |
| Legal Intervention | 1500 | 0.86 | 0.86 | 0.85 | 0.01 |
| Legal Intervention | 2000 | 0.83 | 0.9 | 0.78 | 0.01 |
| Number Nonfatally Shot | 100 | 0.31 | 0.2 | 0.71 | 0.09 |
| Number Nonfatally Shot | 200 | 0.29 | 0.23 | 0.38 | 0.12 |
| Number Nonfatally Shot | 500 | 0.34 | 0.25 | 0.55 | 0.1 |
| Number Nonfatally Shot | 1000 | 0.41 | 0.32 | 0.57 | 0.09 |
| Number Nonfatally Shot | 1500 | 0.64 | 0.54 | 0.79 | 0.04 |
| Number Nonfatally Shot | 2000 | 0.66 | 0.62 | 0.72 | 0.05 |

| | | | | | |
|---|---|---|---|---|---|
| Victim Injured at Home | 100 | 0.57 | 0.53 | 0.63 | 0.13 |
| Victim Injured at Home | 200 | 0.63 | 0.52 | 0.8 | 0.08 |
| Victim Injured at Home | 500 | 0.71 | 0.64 | 0.81 | 0.07 |
| Victim Injured at Home | 1000 | 0.7 | 0.58 | 0.88 | 0.05 |
| Victim Injured at Home | 1500 | 0.75 | 0.66 | 0.87 | 0.05 |
| Victim Injured at Home | 2000 | 0.74 | 0.67 | 0.82 | 0.06 |

Notes: Each numeric cell contains performance (F1, precision, recall) for each model outcome by the proportion of positive class cases that were included in the training data. In Column 2, the proportion of the training data that was a positive class case is listed, with the exception being “random sample” which denotes training data that is randomly sampled up to n=1000 cases and is presented for reference. The train proportion increases the proportion of positive class cases beyond the random sample. The exception is the outcome Victim Injured at Home which omits the train proportion at 10 and 20 percent because the randomly sampled proportion exceeds 20 percent.
